# Investigating Attitudes, Motivations and Key Influencers for vaccine uptake among late adopters of COVID-19 vaccination in Africa

**DOI:** 10.1101/2022.04.20.22274081

**Authors:** A Tariro Makadzange, Charles Lau, Janan Dietrich, Admire Hlupeni, Nellie Myburgh, Patricia Gundidza, Nyasha Elose, Shabir Mahdi, Wilmot James, Larry Stanberry, Chiratidzo Ndhlovu

## Abstract

**Background:** The rapid development of vaccines in response to the COVID-19 pandemic has provided an effective tool for the management of COVID-19. However, in Africa there has been a poor uptake of COVID-19 vaccines with only 15% vaccine coverage compared to the WHO global target of 70%. One of the important drivers has been vaccine hesitancy, understanding late adopters of vaccination can provide insights into the attitudes, motivations and influences that can enhance vaccine uptake.

**Methods:** Between January 4 – February 11, 2022, we conducted a survey among adults presenting for their first dose of a COVID-19 vaccine almost 12-months after the vaccination program began. Vaccines were free and provided at clinics and outreach centers in Harare, Zimbabwe. The questionnaire assessed environmental and individual factors (attitudes, barriers, motivations, key influencers, and information sources) that influenced the decision to present for vaccination. Baseline socio-demographic data and responses to survey questions were summarized using descriptive statistics. Binary logistic regression models were developed to understand factors associated with vaccine confidence.

**Results:** 1016 adults were enrolled into the study, 508 (50%) were female, 126 (12.4%) had HIV co-infection. The median age was 30 years (IQR 22 – 39). Women were more likely to have negative views about the COVID-19 vaccine compared to men (OR 1.51 (95%CI 1.16, 1.97, p=0.002). Women compared to men and older adults (≥ 40 years) compared with youth (18-25 years) were more likely to have ‘major concerns’ about vaccines. Most concerns were about safety with 602 (59.3%) concerned about immediate and 520 (51.2%) about long-term health effects of vaccines. People living with HIV (PLWH) were more likely to perceive vaccines as safe (OR 1.71 (95%CI: 1.07, 2.74, p=0.025), effective (1.68 (95%CI: 1.07, 2.64, p=0.026) and to trust regulatory systems for approving vaccines (OR 1.79 (95% CI: 1.11, 2.89, p=0.017) compared to those without HIV. Internet users were less likely to perceive vaccines as safe (OR 0.72 (95% CI: 0.55, 0.95, p=0.021), effective (OR 0.61 (95% CI: 0.47, 0.80, p<0.001) or trust regulatory processes for approving vaccines (OR 0.64 (95% CI: 0.48, 0.85, p=0.002) compared to non-internet users. Social influence was a key factor in the decision to be vaccinated with family members being the primary key influencers for 560 (55.2%) participants. The most important reason for receiving the COVID-19 vaccine today for 715 (70.4%) participants was the protection of individual health. The most trusted source of information regarding the vaccine was the Ministry of Health (79.7%) and the radio, television and social media were the preferred sources for obtaining this information. Social media was a more likely source for youth and those with higher levels of education.

**Conclusion:** Improving vaccine coverage will need targeted communication strategies that address negative perceptions of vaccines and associated safety and effectiveness concerns. Leveraging normative behavior as a social motivator for vaccination will be important as close social networks are key influences of vaccination. Traditional media remains important for health communication in Africa and should be strengthened to counter social media-based misinformation that drives concerns about safety and effectiveness particularly among internet users.

## Introduction

The severe acute respiratory syndrome coronavirus 2 (SARS-CoV-2) has had a devastating impact on health and socio-economic well-being for millions of people. Since the start of the pandemic there have been over 460 million cases and six million deaths(1). Vaccines are one of the most effective approaches for the public health management of many infectious diseases. In response to the SARS-CoV-2 pandemic, over 300 vaccines have gone into development across various platforms(2). Among these 35 have been approved by at least one country and 10 have been approved by the World Health Organization (WHO)(3). In addition to manufacturing and distribution, widespread vaccination acceptance will be required to achieve sufficient vaccination coverage rates(4).

In pre-pandemic 2019, the WHO identified vaccine hesitancy as one of the top 10 threats to human health (5).Vaccine hesitancy in Africa is poorly understood particularly for adult vaccines and within key subpopulation such as people living with HIV (PLWH)(6). Studies conducted largely prior to the availability of COVID-19 vaccines suggested vaccine acceptance rates would be high in Africa(7). However, as vaccine access improves it is increasingly becoming evident that vaccine hesitancy is a major driver of low COVID-19 vaccine coverage rates in Africa(8). Vaccine acceptance lies on a spectrum and includes early adopters of all vaccines; the vaccine hesitant who take a ‘wait and see’ approach and may present for vaccination at later stages of the vaccine campaign; and those that do not accept all vaccines. Very little is understood about the late adopters that take a ‘wait and see’ approach. These individuals are among those for whom targeted approaches are likely to have the most impact in driving up vaccine coverage rates.

The WHO has set a target of achieving 70% vaccine coverage globally by mid-2022(9). However, Africa currently has less than 15% vaccine coverage with vaccine hesitancy being an important driver of low coverage rates(10). One year after initiating its vaccination program, first dose coverage in Zimbabwe is 45.5% and second dose coverage is 35.4% (MOHCC, Zimbabwe). Over the last several months a plateau in vaccine uptake rates has been observed making the country like many African countries unlikely to reach the WHO target (8). We conducted a survey among individuals who were presenting for their first COVID-19 vaccine at public vaccination centers in Harare, Zimbabwe. The participants were presenting for vaccination one year after initiation of the vaccination program and are late adopters who lie on the vaccine hesitancy spectrum as the ‘wait and seers’(11). The aim of this study was to measure the environmental and individual factors (attitudes, barriers, motivations, key influencers, and information sources) that influence vaccine uptake among late adopters (12).

## Materials and Methods

### Study design

Between January 4^th^ and February 11^th^, 2022, we conducted a quantitative survey through face-to-face interviews.

### Study Sites and Sampling

Eligible adults (≥18 years) who were receiving their first dose of COVID-19 vaccines at public sites providing free vaccine services at 4 City of Harare clinics (Wilkins, Kuwadzana, Budiriro, Mabvuku) including their affiliated outreach sites (Mabelreign, Avondale, and Dzivarasekwa) were consecutively enrolled into the study. All individuals receiving their first COVID-19 vaccine were eligible to participate except those who had obvious cognitive impairments or were unable to provide informed consent.

### Study Procedures

Participants at each site who were receiving the first dose of the vaccine were informed about the study while queueing to receive the vaccine. Those that agreed and were eligible participated in the survey after receiving their vaccination. Participants were reimbursed $5 for participating in the survey.

### Study Measures

A comprehensive questionnaire was prepared based on a user-centered design framework (13). The questionnaire consisted of six main sections including demographics (sex, age, education, socio-economic status), attitudes and views towards COVID-19 vaccines, barriers, motivations, information sources and vaccination experience (Table 1). Age was categorized into 3 groups: ages 18-25 as youth, 26-39 years as young adults and ≥ 40 years as older adults. Vaccine confidence was measured through survey statements related to individual perception on safety, effectiveness of vaccines and confidence in country’s regulatory approval process for vaccines (14) (Table 1). Socioeconomic status was based on an assessment of available resources – electricity, television, refrigerator, bed, battery or generator for power. High socioeconomic status was defined by owning both a battery or a generator for power and a refrigerator, low economic status was defined by neither owning a refrigerator or alternative source of power. In addition, internet usage in the last 30 days, was assessed.

**Table 1.** Survey questions addressing Vaccine convenience and confidence. (Full study questionnaire included in appendix)

| Question | Response options |
| --- | --- |
| **Vaccine convenience** |  |
| How long did it take you to travel to this vaccination site today? | Less than 15 minutes; 15-30 minutes; 31-60 minutes; 1-2 hours; More than 2 hours; Don't know; Prefer not to answer |
| From the time you arrived at the vaccination site, how long did it take you to get your vaccine shot? | Less than 10 minutes; 10-20 minutes; 20-30 minutes; 30-60 minutes; More than 60 minutes; Not applicable |
| How easy or difficult was it to register for a vaccination appointment? | Very easy, Somewhat easy; Neither easy nor difficult; Somewhat difficult; Very difficult |
| How easy or difficult was it to find a vaccination site? | Very easy, Somewhat easy; Neither easy nor difficult; Somewhat difficult; Very difficult |
| How easy or difficult was it to get to the vaccination site? | Very easy, Somewhat easy; Neither easy nor difficult; Somewhat difficult; Very difficult |
| How easy or difficult was it to financially afford transportation to the vaccination site? | Very easy, Somewhat easy; Neither easy nor difficult; Somewhat difficult; Very difficult |
| How easy or difficult was it to find a vaccination site with convenient hours? | Very easy, Somewhat easy; Neither easy nor difficult; Somewhat difficult; Very difficult |
| Did you need to do any of the following in order to get vaccinated today? Please select all that apply | Arrange for childcare; Take time off from paid work; Earn less money because you weren't working; None of the above |
| **Vaccine Confidence** |  |
| When the COVID-19 vaccines first became available, what was your view towards COVID-19 vaccines? | Very positive; Somewhat positive; Neutral; Somewhat negative; Very negative |
| It is common for people to be hesitant or unsure about getting vaccinated. Did you ever have concerns about getting the COVID-19 vaccine? | Yes, major concerns; Yes, minor concerns; No concerns |
| Why were you concerned about getting a COVID-19 vaccine? Please select all that apply | Whether the vaccine has been tested enough; The vaccine is still new; Effectiveness of the vaccine; Immediate side effects; Long-term health effects; Don't see need for vaccine; Other (specify); None of the above |
| In general, COVID-19 vaccines are safe | Strongly agree; Somewhat agree; Neutral; Somewhat disagree; Strongly disagree; Don't know; Prefer not to answer |
| I am confident that my country's regulation process approved the COVID-19 vaccine, only when it was shown to be safe | Strongly agree; Somewhat agree; Neutral; Somewhat disagree; Strongly disagree; Don't know; Prefer not to answer |
| I am confident that COVID-19 vaccines are effective in preventing the disease. | Strongly agree; Somewhat agree; Neutral; Somewhat disagree; Strongly disagree; Don't know; Prefer not to answer |
| **Motivators/Influencers and Reasons for vaccination today** |  |
| Which of the following people help to convince you to get vaccinated, if any? Please select all that apply. | Spouse; Children; other family member; Healthcare worker; Religious leader; Community leader; Co-worker; Media; Other (specify); None of the above |
| Why did you get vaccinated today? Please select all that apply. | To protect my health; To protect health of my family; To protect health of people in community; To get back to work or school; To resume social activities; To resume travel; Because other people encouraged me to get vaccinated; Other (specify); Don't know; Prefer not to answer |
| What is the most important reason you got vaccinated today? Select one reason only. | To protect my health; To protect health of my family; To protect health of people in community; To get back to work or school; To resume social activities; To resume travel; Because other people encouraged me to get vaccinated; Other (specify); Don't know; Prefer not to answer |
| Do you agree or disagree with the following statement: I came to get vaccinated because people important to me encouraged me to get vaccinated | Strongly agree; Somewhat agree; Neutral; Somewhat disagree; Strongly disagree |
| Do you agree or disagree with the following statement: I came to get vaccinated because people important to me got a COVID-19 vaccine themselves? | Strongly agree; Somewhat agree; Neutral; Somewhat disagree; Strongly disagree |
| Information Sources |  |
| In the past 30 days, who have you received information from about COVID-19? Select all that apply. | Ministry of health; Local health authorities; World Health Organization (WHO); Pharmaceutical companies producing the COVID-19 vaccines; Health scientists and researchers; Your doctor; Your local clinic; Other healthcare professionals like nurses or pharmacists; Alternative health providers (e.g., faith or traditional healer; Friends or family; Religious leaders; Other people's experience or knowledge; Other (specify); None of the above |
| What are your top 3 sources of trusted information about COVID-19 vaccines? | Ministry of health; Local health authorities; World Health Organization (WHO); Pharmaceutical companies producing the COVID-19 vaccines; Health scientists and researchers; Your doctor; Your local clinic; Other healthcare professionals like nurses or pharmacists; Alternative health providers (e.g., faith or traditional healer; Friends or family; Religious leaders; Other people's experience or knowledge; Other (specify); None of the above |
| In the past 30 days, where have you obtained information about COVID-19 vaccines? Select all that apply | Radio; Television; WhatsApp; Facebook; TikTok; Instagram; Other social media; Other (specify) |

### Statistical analysis

Data were analyzed using Stata version 13 (College Station, Texas 77845 USA). Baseline socio-demographic data and responses to survey questions were summarized using means and standard deviations (SD) for normally distributed data or medians and interquartile ranges (IQRs) for non-parametric continuous variables. Absolute numbers, proportions and percentages were used for categorical data. The chi-squared test was used to test for significance between outcome variables and demographic variables such as gender, age, education, economic status, previous personal experience with COVID-19, HIV status and internet use over the last 30 days.

Binary logistic regression models were developed to examine factors associated with ‘very negative’ and ‘somewhat negative’ views to vaccines when they were first available, as well as concerns about vaccines. The outcome variables were dichotomized (Very negative/somewhat negative or Neutral/somewhat positive/very positive; Major concerns/Minor concerns or No concerns; Strongly agree or somewhat agree/neutral/somewhat disagree/strongly disagree). The variables evaluated included demographic factors such as gender, age, education, and income (a proxy for which was defined by availability of alternate sources of energy (battery or generator), and/or refrigerator ownership), personal experience with COVID-19 – knowing someone who became severely ill or died from COVID-19, internet usage and HIV status. P-values were two-sided.

### Ethics Approvals

The protocol, consent forms and recruitment materials were reviewed and approved by the Medical Research Council of Zimbabwe, Joint Research Ethics Committee (JREC) of Parirenyatwa Hospital, the University of Zimbabwe, and Harare City Health Departments prior to initiation of the study. All amendments to the protocol, consent forms and/or recruitment materials were approved by the institutional review boards before they were implemented. All participants provided written informed consent.

## Results

A total of 1016 adults were enrolled into the study; 1013 (99.7%) received the Sinopharm vaccine and 3 (0.3%) received the Sinovac vaccine. Gender was equally distributed in the cohort with 508 (50%) females, and a median age of 30 years (IQR 22-39) (Table 2). All participants were of African descent and most had attained some level of high school education. The self-reported comorbid conditions were diabetes (1.5%), cardiac disease (0.4), respiratory illnesses (2.3%), hypertension (6.6%). HIV positive status was reported by 126 (12.4%) participants. The study participants were urban residents with high levels of electricity use (87.7%), refrigerator (75.8%) and television (88%) ownership. Having a ‘battery or generator for power’ was reported by 240 (23.6%) participants. Ownership of these resources was used to classify participants as high, middle, or low socioeconomic status (Table 1). Among study participants 428 (42.1%) reported knowing someone who was seriously ill or died from COVID-19 disease; these were close relatives (e.g., siblings, spouse or parents) in 30% of respondents. Internet use in the past 30 days was reported by 420 (41.4%) participants (Table2).

**Table 2.** Baseline cohort demographics.

| Characteristic | N(%) |
| --- | --- |
| **Gender (Female)** | 508 (50%) |
| **Median age (IQR)** | 30 (22-39) |
| **Age groups** |  |
| 18-25 (Youth) | 368 (36.2%) |
| 26-39 (Young adults) | 409 (40.3%) |
| ≥40 (Older adults) | 239 (23.5%) |
| **Ethnicity (Black African)** | 1016 (100%) |
| **Highest Level of Education** |  |
| No formal schooling | 5 (0.5%) |
| Primary school<sup>1</sup> | 84 (8.3%) |
| Lower Secondary school<sup>2</sup> | 735 (72.4%) |
| Higher Secondary<sup>3</sup> | 117 (11.5%) |
| Tertiary Education | 74 (7.3%) |
| **Co-morbid conditions** |  |
| Diabetes | 15 (1.5%) |
| Cardiac Disease | 4 (0.4%) |
| Respiratory Illness | 23 (2.3%) |
| Hypertension | 67 (6.6%) |
| HIV | 126 (12.4%) |
| **Socioeconomic status** |  |
| High<sup>4</sup> | 172 (16.9%) |
| Middle<sup>5</sup> | 598 (58.9%) |
| Low<sup>6</sup> | 246 (24.2%) |
| **Internet use** |  |
| In the past 30 days have you used the internet? | 420 (41.4%) |
| **Personal Covid experience** |  |
| Know someone who became seriously ill or died as a result of Covid | 428 (42.1%) |

<sup>1</sup> Primary up to 7<sup>th</sup> grade
<sup>2</sup> High school completing up to and including Ordinary Level (GSCE or 11<sup>th</sup> Grade)
<sup>3</sup> Higher secondary completing Advanced Level (or 13<sup>th</sup> Grade)
<sup>4</sup> High socioeconomic status defined by owning a generator or battery as an alternate source of power and a refrigerator
<sup>5</sup> Middle socioeconomic status owned one but not the other
<sup>6</sup> Low socioeconomic status neither owned a refrigerator or a generator

### Vaccine convenience

Time to travel to the vaccination site was <15 minutes for 443 (43.6%) participants. Time taken to receive the shot after arrival was less than 10 minutes for 338 (33.3%) and less than an hour for 839 (82.6%). Appointments were not needed to receive the vaccine. Finding a vaccination site was described as ‘very easy’ by 853 (84.2%), it was ‘very easy’ to get to the vaccination center for 889 (87.7%), it was ‘very easy’ to financially afford transportation to the site for 877 (86.5%) and ‘very easy’ to find a vaccination site with convenient hours for 786 (77.5%) participants. Among the participants 106 (10.5%) indicated that they had to arrange for childcare, 207 (20.4%) had to take time off from paid work and 112 (11%) earned less money because they were not working and receiving the vaccine.

### Vaccine confidence

When asked “When the COVID-19 vaccines first became available, what was your view towards COVID-19 vaccines?”, 477(47%) had ‘very negative views’ (Figure 1) and 198 (19.5%) had ‘somewhat negative’ views. Women were more likely than men (OR 1.51 (95% CI: 1.16,1.97, p=0.002)) and young adults were more likely than youth (OR 1.37 (95%CI: 1.01, 1.86, p=0.043)) to have ‘very negative’ or ‘somewhat negative’ views towards COVID-19 vaccines (Table 3).

**Table 3.**
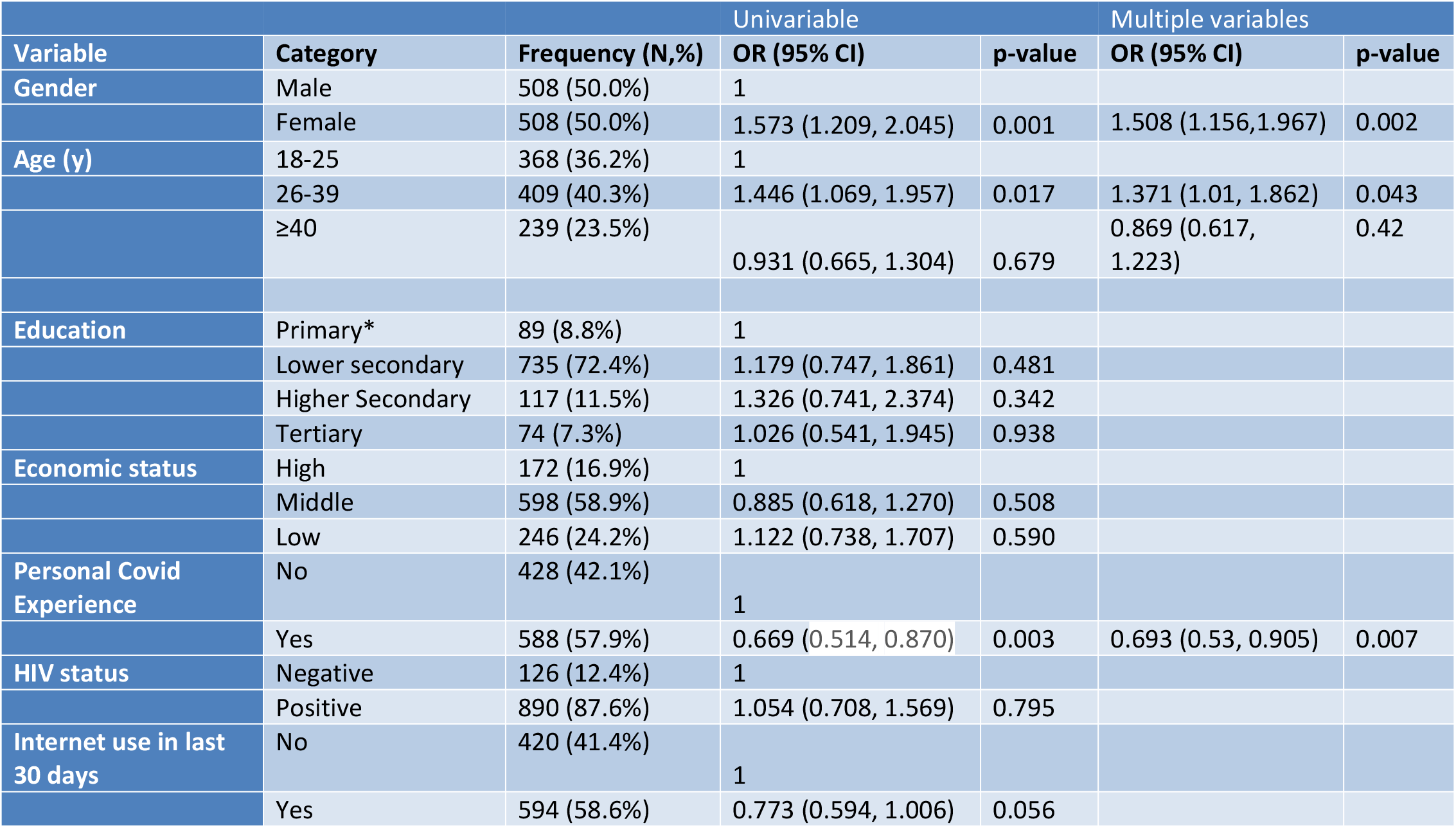
Factors associated with having a Very negative or somewhat negative view towards COVID-19 vaccines when they first became available.

**Figure 1.**
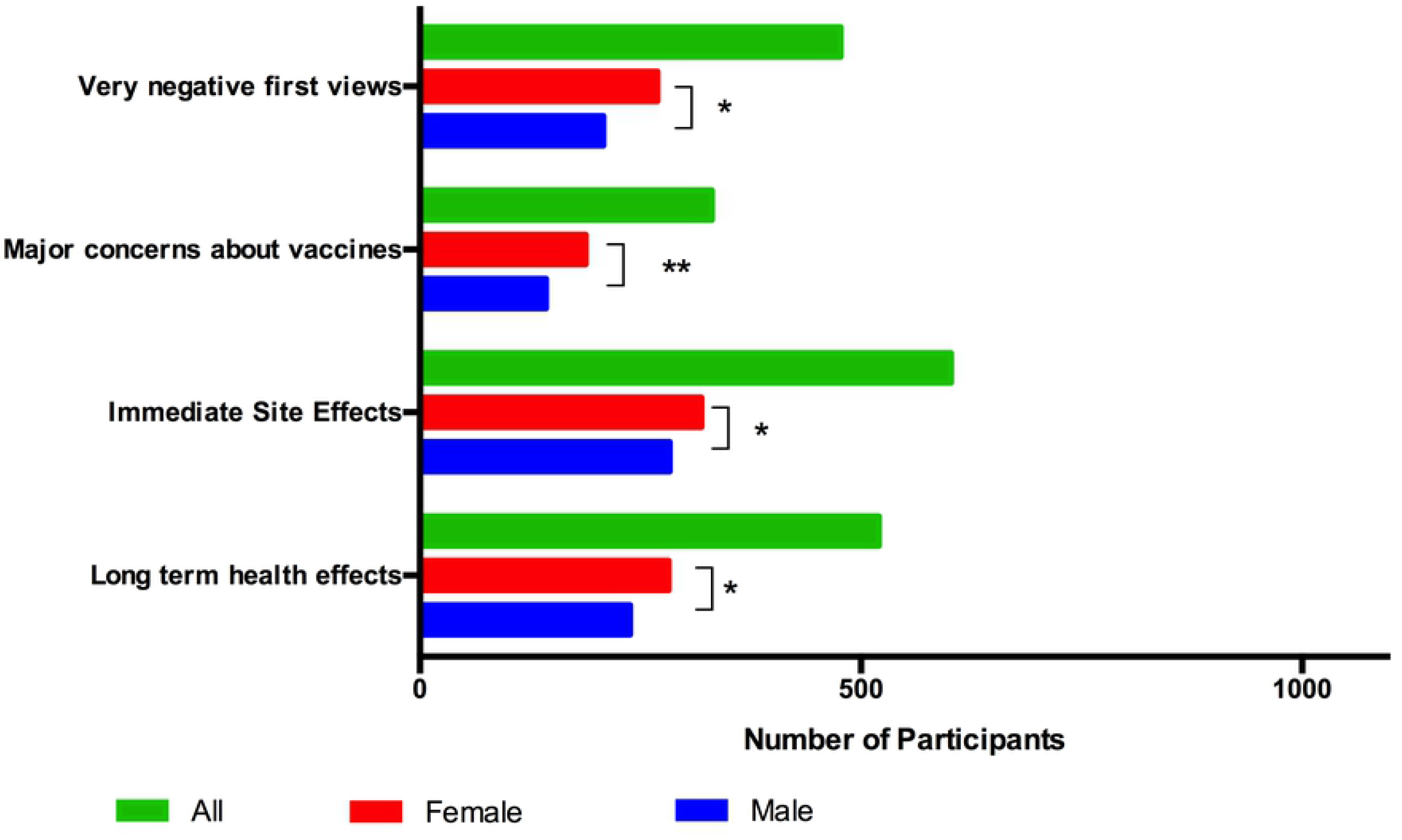
Attitudes towards COVID-19 vaccine and Major concerns. Participants were asked “When vaccines first became available, what was your view towards COVID-19 vaccines?” and “It is common for people to be hesitant or unsure about getting vaccinated. Did you ever have concerns about getting the COVID-19 vaccine?”. Women had more negative views when vaccines were first available, more major concerns and concerns about side effects (* p<0.05; ** p<0.001)

A third of participants (32.6%) had major concerns about receiving the COVID-19 vaccine (Figure 1). Gender, age and HIV status were associated with differences in major concerns. Women were more likely than men (OR 1.5 (95%CI: 1.15, 1.95), p=0.003) to have major concerns (Table 4). Young adults (OR 1.52 (95% CI: 1.11, 2.09, p=0.009) and older adults (OR 2.46 (95%CI: 1.74, 3.49, p<0.001) were more likely than youth (Table 4). Similarly, people living with HIV (PLWH) were almost twice as likely as those without HIV (OR 1.72 (95%CI: 1.18, 2.52, p=0.005) to have major concerns about COVID-19 vaccines (Table 4). Individuals who knew someone who became seriously ill or died because of COVID-19 were less likely to have negative views (OR 0.69 (95%CI: 0.53-0.91, p=0.007)) or major concerns (OR 0.69 (95%CI: 0.52-0.91, p=0.009)) compared to those who did not (Tables 3 and 4).

**Table 4.** Major concerns about COVID-19 vaccine, Immediate side effects and Long-term health effects

| Variable | Category | Frequency | Major Concerns |  | Immediate Side Effects |  | Long term Health Effect |  |
| --- | --- | --- | --- | --- | --- | --- | --- | --- |
|  |  |  | OR (95% CI) | p-value | OR (95% CI) | p-value | OR (95% CI) | p-value |
| Gender | Male | 508 | 1 |  | 1 |  | 1 |  |
|  | Female | 508 | 1.5 (1.151, 1.953) | 0.003* | 1.342 (1.044, 1.725) | 0.022* | 1.416 (1.106, 1.812) | 0.006* |
| Age (y) | 18-25 | 368 | 1 |  | 1 |  | 1 |  |
|  | 26-39 | 409 | 1.522 (1.111, 2.085) | 0.009* | 1.48 (1.109, 1.975) | 0.008* | 1.608 (1.211, 2.135) | 0.001* |
|  | ≥40 | 239 | 2.462 (1.737, 3.489) | <0.001* | 1.055 (0.76, 1.465) | 0.749 | 1.142 (0.824, 1.582) | 0.426 |
| Education | Primary | 89 | 1 |  | 1 |  | 1 |  |
|  | Lower secondary | 735 | 0.698 (0.444, 1.098) | 0.12 | 0.949 (0.603, 1.491) | 0.819 | 1.172 (0.754, 1.821) | 0.48 |
|  | Higher Secondary | 117 | 0.742 (0.418, 1.319) | 0.309 | 0.773 (0.441, 1.355) | 0.369 | 1.219 (0.702, 2.117) | 0.482 |
|  | Tertiary | 74 | 0.994 (0.529, 1.870) | 0.986 | 0.586 (0.313, 1.094) | 0.093 | 1.317(0.709, 2.443) | 0.383 |
| Economic status | High | 172 | 1 |  | 1 |  | 1 |  |
|  | Middle | 598 | 0.846 (0.59, 1.215) | 0.365 | 1.075 (0.709, 1.560) | 0.803 | 1.28 (0.866, 1.891) | 0.215 |
|  | Low | 246 | 1.195 (0.795, 1.796) | 0.393 | 0.905 (0.642, 1.278) | 0.572 | 1.016 (0.723, 1.427) | 0.926 |
| Personal Covid Experience | No | 428 | 1 |  | 1 |  | 1 |  |
|  | Yes | 588 | 0.633 (0.482, 0.830) | 0.001* | 0.852 (0.661, 1.097) | 0.215 | 0.688 (0.536, 0.884) | 0.003* |
| HIV status | Negative | 126 | 1 |  | 1 |  | 1 |  |
|  | Positive | 890 | 1.723 (1.179, 2.519) | 0.005* | 1.558 (1.047, 2.23) | 0.029* | 1.527 (1.044, 2.233) | 0.029* |
| Internet use in last 30 days | No | 420 | 1 |  | 1 |  | 1 |  |
|  | Yes | 594 | 1.036 (0.794, 1.362) | 0.796 | 0.778 (0.604, 1.003) | 0.053 | 1.158 (0.902, 1.488) | 0.25 |

Immediate health concerns were a concern for 602 (59.3%) and long-term health effects for 520, (51.2%) (Figure 1). Young adults compared to youth (OR 1.48 (95%CI: 1.11, 1.98), p=0.008) and PLWH (OR 1.56 (95%CI: 1.05-2.23, p=0.029) compared to those without were more likely to express concerns about immediate side effects (Table 4). Long-term health effects were a particular concern for women compared to men, and young adults compared with youth (Table 4). Those with personal knowledge of someone who was seriously ill or died from COVID-19 were less likely to indicate concerns about long-term health effects (OR 0.69 (95% CI: 0.54, 0.88, p=0.003) (Table 4).

The concern that ‘the vaccine has not been tested enough’ was expressed by 223 (21.9%), that the ‘vaccine was still new’ by 190 (18.7%) and 167 (16.4%) indicated that they did ‘not see a need for a vaccine’. Among the respondents, 293 (28.8%) chose ‘other’ for their concerns. The 293 participants’ concerns focused on fear of death, infertility, health effects of vaccination, interaction with medications and comorbid conditions and interference with pregnancy and breastfeeding as well as conspiracy theories (Table 5).

**Table 5.**
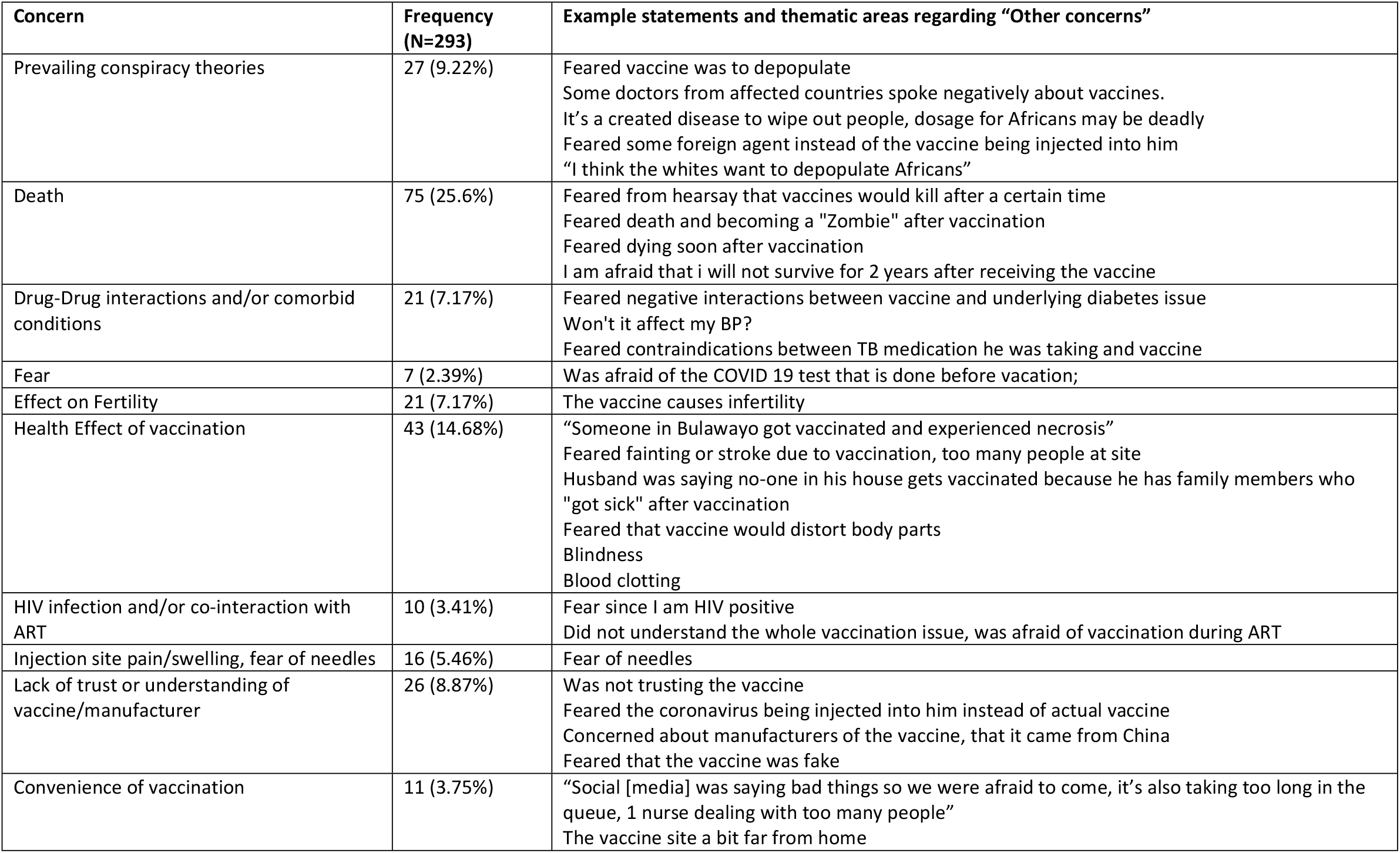
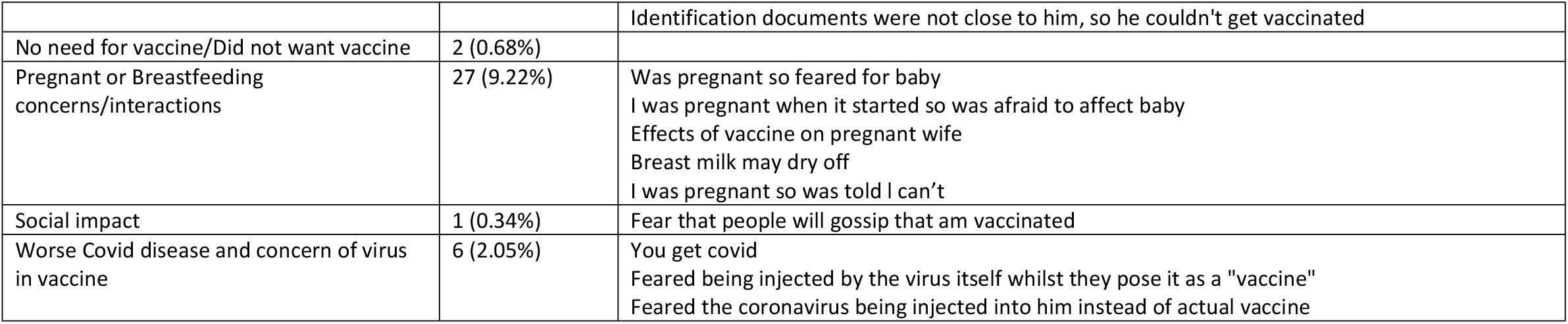
Summary of primary themes arising among the concerns expressed by study participants who listed concerns as “other”

Perceived vaccine safety was high with 728 (71.5%) participants strongly agreeing with the statement “In general COVID-19 vaccines are safe” (Figure 2). The odds of strongly agreeing that vaccines are safe decreased with education and internet use (Suppl Table 1). PLWH had higher odds of perceiving vaccines as safe compared to those without HIV (OR 1.71 (95%CI: 1.07, 2.74, p=0.025) (S1 Table). Although most respondents felt that the Sinovac/Sinopharm vaccine was either very safe (500 (49.2) or somewhat safe (175 (17.2%)), 311 (30.6%) indicated that they did not know if the vaccine that had received was safe.

**Figure 2.**
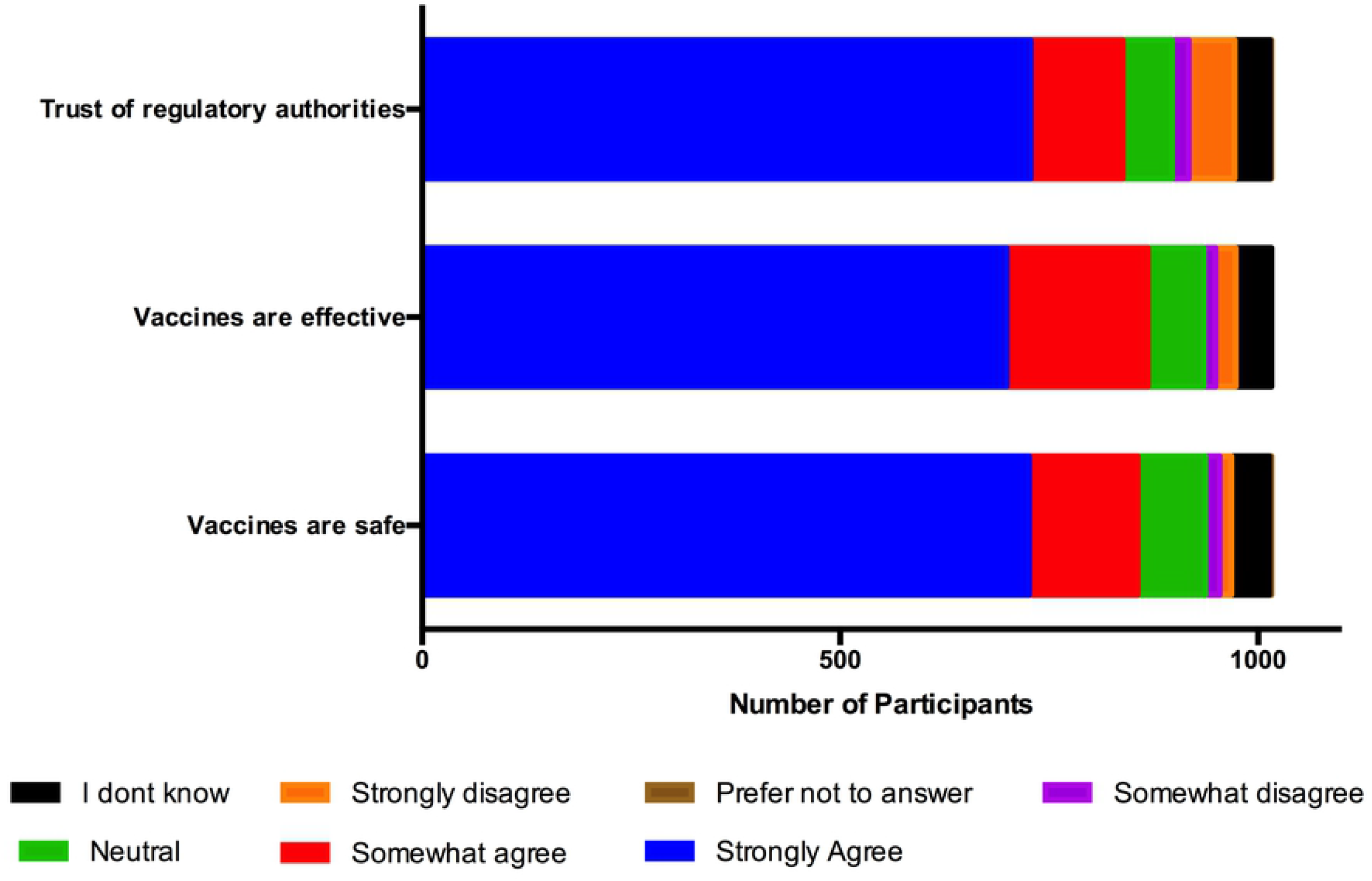
Perceived safety, effectiveness, and Trust in regulatory authorities. Participants were asked “In general COVID-19 vaccines are safe”; “I am confident that COVID-19 vaccines are effective in preventing the disease” “I am confident that my country’s regulatory process approved the COVID-19 vaccine only when it was shown to be safe”.

Perceived vaccine effectiveness was high with 699 (68.8%) participants strongly agreeing with the statement “I am confident that COVID-19 vaccines are effective in preventing the disease” (Figure 2). The odds that one perceived vaccine to be effective in preventing disease decreased with increasing education and with internet use (S2 Table). PLWH compared to those without HIV (OR 1.68 (95%CI: 1.07, 2.64) and those with a personal experience with COVID-19 compared to those without (OR 1.47 (95%CI: 1.12, 1.95, p=0.006) had higher odds to strongly agree that COVID-19 vaccines were effective in preventing disease (S2 Table)

Perceived confidence in national regulatory processes was high with 728 (71.7%) strongly agreeing with the statement “I am confident that my country’s regulatory process approved the COVID-19 vaccine only when it was shown to be safe” (Figure 2). Increasing levels of education and internet use were associated with lower odds of strongly agreeing with the statement (S3 Table). Participants with a personal experience with COVID-19 (OR 1.37 (95%CI: 1.03, 1.82), p=0.028) were more likely to be confident in national regulatory processes. Similarly, PLWH were more likely than those without HIV (OR 1.79 (95%CI: 1.11, 2.89), p=0.017) to strongly agree with the statement (S3 Table).

Internet users consistently had lower perceived confidence in vaccine safety (OR 0.72 (95%CI: 0.55, 0.95), p=0.02), lower perceived confidence in vaccine effectiveness (OR 0.61 (95%CI: 0.47, 0.50, p<0.001) and lower perceived confidence in regulatory processes (OR 0.64 (95%CI: 0.48, 0.85, p=0.002) than those that had not used the internet in the last 30-days (S1-3 Table).

### Key Influencers

Normative behavior can be a key motivator of vaccine uptake. The influence of others was important with 684 (67.3%) participants indicating that they strongly agreed with the statement “I came to get vaccinated because people important to me encouraged me to be vaccinated”. In addition, 702 (69.1%) of respondents strongly agreed with the statement “I came to get vaccinated today because people important to me got a COVID-19 vaccine” (Figure 3a). For 561 (55.2%) participants family members were the key influencers. Religious leaders (98, (9.7%)), community leaders (56, (5.5%)), healthcare workers (133, (13.1%)) and co-workers (137 (13.5%)) were less commonly listed as key influencers (figure 3b).

**Figure 3.**
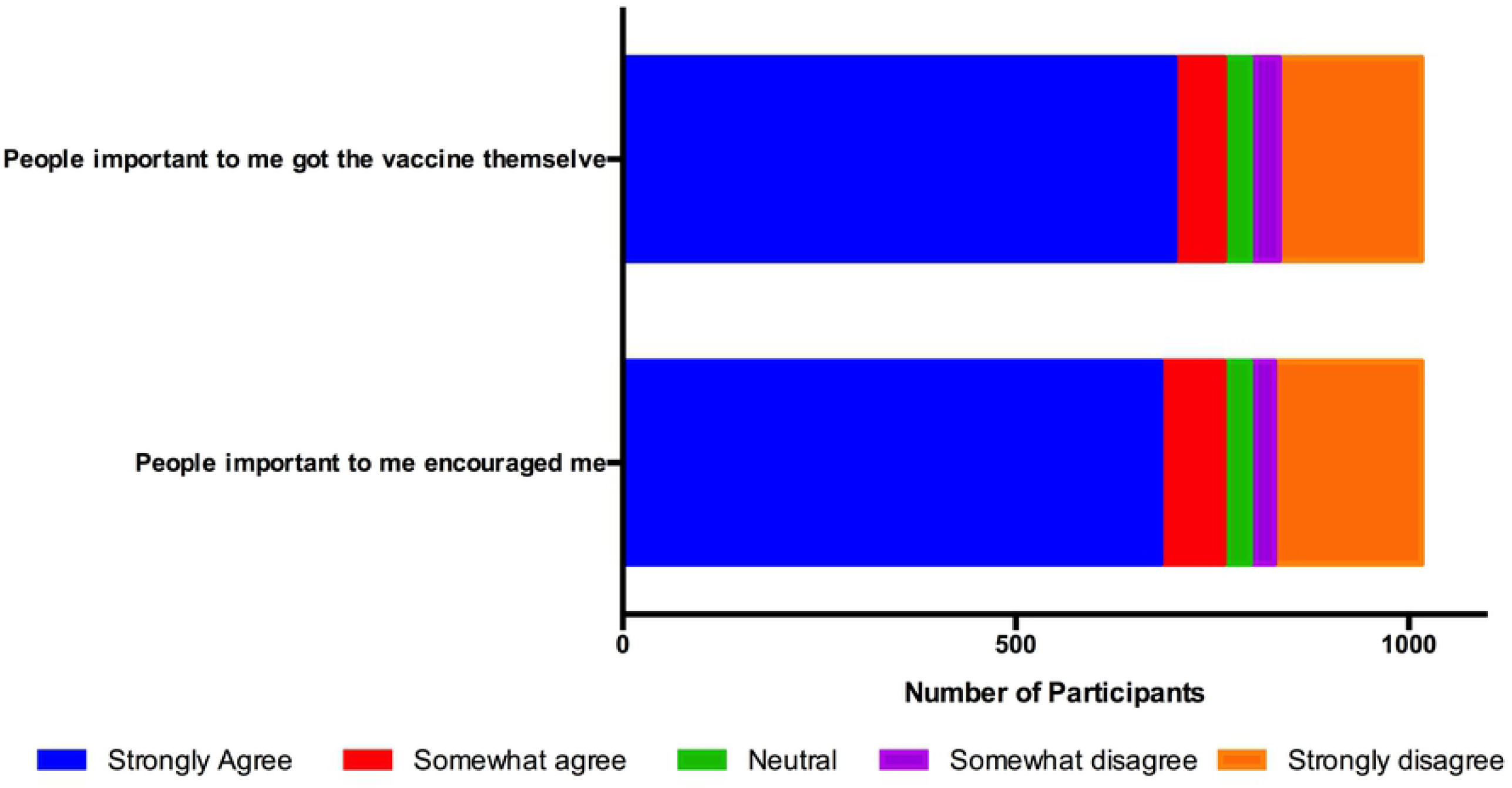
Social influence. **(a)** Participants indicated if they agreed with the statements: “I came to get vaccinated because people important to me encouraged me to be vaccinated” and “I came to get vaccinated today because people important to me got a COVID-19 vaccine” **(b)** Participant responses when asked “Which of the following help to convince you to get vaccinated, if any?”

The reason for getting vaccinated today was to ‘protect my health’ for 862 (84.8%), to ‘protect the health of my family’ for 590 (58.1%), ‘to protect health of the people in the community’ for 552 (54.3%) and ‘to get back to work or school’ for 451 (44.7%) (Suppl Fig. 7). When asked the *single* most important reason for getting vaccinated today the top two most important reasons were ‘To protect my health’ for 716 (70.4%) participants, and to ‘get back to work or school’ for 152 (14.9%) (Figure 4). Among PLWH, protection of the health of the family (68.3% vs. 56.6%, p<0.001), and the health of the community (65.1% vs 52.8%, p<0.001) were more frequently cited as reasons for vaccination compared to those without HIV infection. Resumption of social activities (2.6%), travel (2.9%) and ‘because others encouraged me to’ (2.3%) were the single most important reason for a very small number of participants (Figure 4).

**Figure 4.**
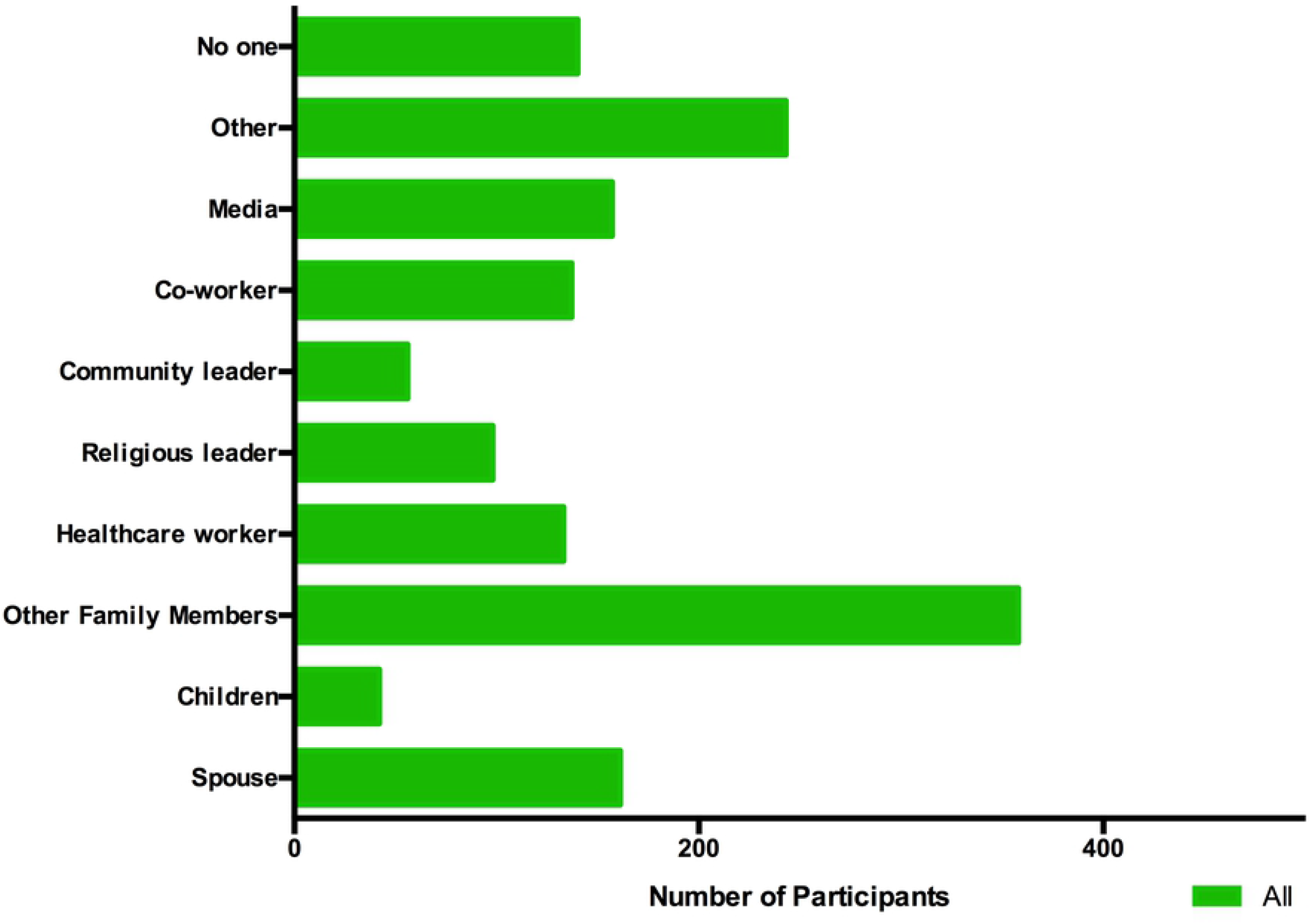
Reasons for presenting for vaccination today among late adopters of vaccination. Participants were asked “Why did you get vaccinated today?”. Multiple responses were allowed (green). The subsequent follow up question was “What is the most important reason you got vaccinate today?”, a single response was allowed.

### Sources of Information

We asked the question ‘In the past 30 days, who have you received information from about COVID-19?’. Friend or family were the most common sources of information for 790, (77.8%), Ministry of Health for 773 (76.1%), local clinic for 596 (58.7%), religious leaders for 463 (45.6%), and the World Health Organization (WHO) for 467 (46%) (Figure 5). When asked the top 3 sources of *trusted* information about COVID-19 vaccines the primary sources were the Ministry of Health (79.7%), WHO (50.8%), local clinic (35%), local health authorities (31.1%), and friends or family (22%) (Figure5). The WHO was a more important source of information for men than women (50.4% vs. 41.5%, p<0.05), whereas women more frequently listed their local clinic (52.2% vs. 65.2%, p<0.0001) and religious leaders (56.3% and 34.8%, p<0.0001) among their top 3 trusted sources for information. Doctors and other health professions were in the top 3 trusted sources for 16.6% and 21.9% respectively, and alternative health providers for 4.4%.

**Figure 5.**
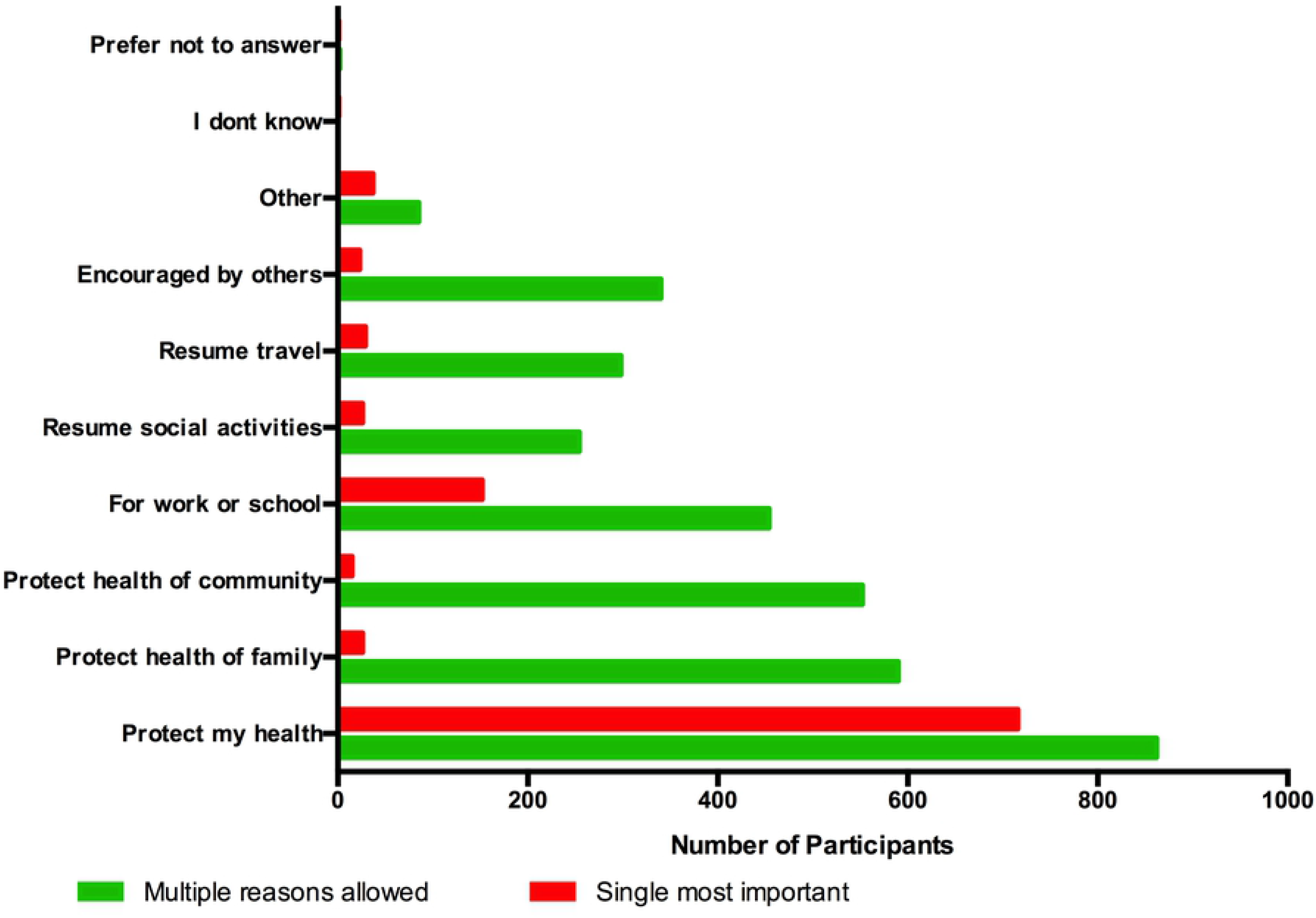
Sources and Trusted sources of information. Participants were asked “In the past 30 days, who have you received information from about COVID-19?” multiple responses were allowed (Green). Participants were subsequently asked “What are your top 3 sources of trusted information about COVID-19 vaccines?” (Red - only 3 responses allowed)

We assessed the use of media as an information source by asking ‘In the past 30 days, where have you obtained information about COVID-19 vaccines?’. The radio (86.5%), television (76.7%), WhatsApp (62.2%) and Facebook (36.9%) were the primary media sources of information (Figure 6). Radio use had high penetration and more likely to be a source for young adults compared with youth (OR 1.85 (95%CI 1.2-2.84), p=0.005) (S4 Table). Although television had high penetration as an information source, its use tracked with education. The odds of receiving your information from television were twice as high in those with lower secondary (OR 2.22 (95%CI 1.39-3.56), p=0.001) and higher secondary education (OR 2.72 (95%CI 1.35-5.48), p=0.005) than those with primary education only (S5 Table). Social media primarily WhatsApp and Facebook as a source of information tracked strongly with age and education. Older adults had much lower odds of obtaining information from WhatsApp (OR 0.41 (95% CI 0.3-0.58), p<0.001) (S6 Table) or Facebook (OR 0.3 (95% CI (0.2-0.45), p<0.001) compared to youth (S7 Table). Similarly, for those with secondary and tertiary education compared to those with primary education (S6 Table).

**Figure 6.**
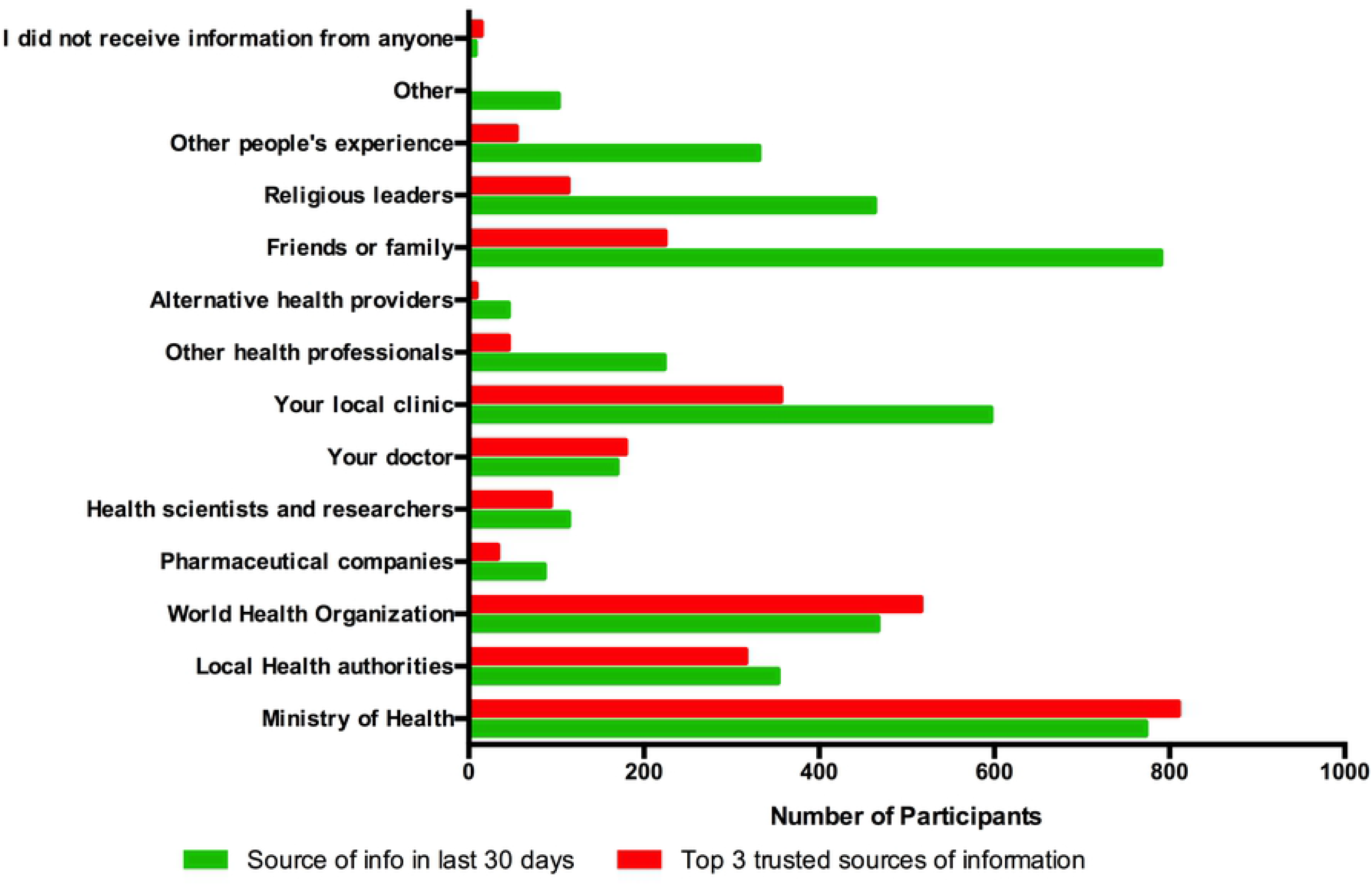
Traditional and social media sources of information. Participants were asked “In the past 30 days, where have you obtained information about COVID-19 vaccines?” and were allowed to select all that applied.

## Discussion

The WHO has set an ambitious goal for COVID-19 vaccination anticipating that over 70% of populations globally will be vaccinated by the end of 2022(9). However, vaccine hesitancy is threatening that goal. The Strategic Advisory Group of Experts (SAGE) on immunization has defined vaccine hesitancy as a behavior influenced by convenience, confidence and complacency towards vaccines(11). Removing practical barriers to vaccination has strengthened various vaccination programs(15-17). Convenience was not a barrier in this study with most participants indicating that the sites were accessible and the service efficient. This may not be the case for most African populations particularly those in rural settings. Improving access is important. In this cohort, we were able to assess individual factors that may influence vaccine uptake when access barriers are minimized.

Vaccine confidence based on perceptions of safety, effectiveness and trust in regulatory processes can influence vaccine uptake. Perceptions of COVID-19 vaccines when they first came out were largely negative in this cohort, likely contributing to the ‘wait and see’ approach of participants. Female gender and increasing age were more likely to be associated with negative perceptions. Gender inequities to vaccination are increasingly recognized as a major risk for successfully achieving full COVID-19 vaccination coverage with several studies suggesting that vaccine hesitancy may be higher in women(18-20). The same was observed in this cohort, with concerns focused primarily on vaccine safety. Unfortunately, up to a third of participants did not know if the vaccine that they received was safe. Communication strategies to enhance vaccine coverage will need to be gender specific and focused on information around the safety of vaccines in general and particular vaccines. The lack of consistent communication on vaccine safety in pregnancy, and ongoing social media misinformation about fertility may be important drivers of delayed vaccine uptake among women in this cohort who were concerned about effects on pregnancy, breast feeding and fertility (21, 22).

Enhancing COVID-19 vaccination uptake among PLWH especially in countries such as Zimbabwe with high HIV seroprevalence (23) will be important. People living with HIV may be at increased risk of hospitalization and death from COVID-19 particularly those that are viremic or have low CD4 counts (24, 25). In addition, immunocompromised individuals, which include people living with uncontrolled HIV infection may be an important source for ongoing SARS-CoV-2 evolution(26). Within this study cohort, PLWH were almost twice as likely to have major concerns about COVID-19 vaccines and side effects than those without HIV. They were concerned about the impact on their health as well as vaccine-drug interactions with their antiretroviral therapy (ART). Vaccination rates among PLWH across different geographics are not fully understood with some studies suggesting that they may at best be like the general population(27, 28). However, vaccination coverage targets for PLWH will likely need to be higher than those for the non-immunocompromised population, particularly in settings where many people are not on ART and with high proportions of individuals with advanced HIV disease (29). Interestingly PLWH were more likely to perceive vaccines as safe and effective and trust government regulatory processes. This may be because PLWH on chronic therapy interact with the healthcare system often and have come to trust in and rely on the services.

Perceptions of safety, effectiveness and trust in regulatory authorities also tracked significantly with education and income status. Those with higher education and better income resources were less likely to perceive the vaccines as effective or trust in regulatory processes. This may be an important trend to watch closely as the more educated and higher income individuals are often key influencers in African families and communities. Ensuring that we design communication strategies that effectively target these groups will be important, particularly as their access of social media-based information and disinformation is also significantly higher. In this study we observed that social influence was a key factor in the motivation to engage in vaccination. The descriptive norms i.e., beliefs about what others are doing regarding the behavior were important motivators. The encouragement of others and the vaccination behaviors of ‘people important to them’ influenced this cohort of late adopters to seek vaccination. Family members were the most important influencers. Similarly, personal experience in knowing someone who was seriously ill or died from COVID-19 also had a significant impact on attitudes towards vaccines. Other key influencers were religious leaders and local clinics particularly for women. Local nurse-led clinics are generally free in Zimbabwe and more accessible than other clinics with other health providers such as medical doctors (30).

This pandemic has shown the importance of media and communications in guiding vaccine uptake. Social media and the anti-vax movement are global and have become an important driver of vaccine hesitancy(22, 31). Politics and trust in governments have been important in driving vaccine uptake in various parts of the world(32). Although levels of trust in government may be low in many countries, we found in this study that trust in government as a source of information on health and specifically COVID-19 was very high. Most participants cited the Ministry of Health as top trusted sources of information. Building and maintaining that trust in Health Ministries and regulatory processes will be critical in ensuring the success of COVID-19 and other vaccination programs.

Traditional media – radio and television were the main media sources of information. Internet based media sources were less commonly cited as major sources with only 41% of the cohort using the internet over the last 30 days. Television and radio are largely state controlled providing the authorities with an opportunity to disseminate accurate information on the safety and effectiveness of vaccines. However social media particularly WhatsApp is growing in importance and was an important source of information particularly for youth, and those with a higher economic status and more education. Communication strategies to enhance vaccine uptake should use both traditional and social media platforms but must be targeted by gender, age on each medium to be most effective.

The study provides valuable insights into late adopters of vaccination in an African setting one year after the national COVID-19 vaccination program began. Because we studied those that made the decision to be vaccinated, this research does not provide insights into those that are continuing to ‘wait and see’. Although our understanding of those that are still on the sidelines of vaccination is not yet enhanced that insights generated from this cohort will enable us to start to effectively target the ‘wait and see’ population.

The study also focused on an urban population where access was not an issue. Further understanding of barriers, motivations, and attitudes among rural populations in Africa will be important. An important limitation is that our survey was not designed to evaluate complacency or hesitancy in-depth. We observed that as many as 16.4% of participants indicated that they ‘Do not see need for vaccine’. Complacency has been described as a major barrier in Africa and may become increasingly important as milder variants of SARS-CoV-2 emerge and seroprevalence from natural infection increases(33). We also did not specifically survey healthcare workers who in some settings may be key drivers of vaccine hesitancy. In sharing this data with healthcare workers many indicated that their own personal knowledge about vaccines was limited, and they would have appreciated further training and vaccine knowledge reinforcement. The impact of healthcare worker hesitancy on program success is critical and needs to be studied and addressed(34, 35).

The study data highlights the need for consistent and targeted information that is gender and age specific and targeted to subpopulations such as PLWH. Information must be delivered by trusted sources such as the Ministry of Health via traditional media platforms such as radio and television that are heavily accessed and can be used to address misinformation delivered on social media. The vaccination encounter should equip friends and family with accurate information on specific vaccines and side effects so they can act as informed social influencers for their family and friends. These individuals can be key ambassadors for vaccination within their social network. In addition, educating religious leaders who have influence over their congregations and are key sources of information, particularly among women, will be important. Vaccine hesitancy can only be tackled effectively by taking on a targeted approach to health communications that builds trust within communities using trusted communicators.

## Data Availability

All relevant data are within the manuscript and its Supporting Information files.

## Acknowledgements

The study would like to acknowledge all members of the study team. Mrs Norest Beta, Mr Taddy Mwarumba, Mr Edward Makaha, Mr John Musvibe, Ms Ruvimbo Chigumbu, Mr Jaffta Chikwonoworo, Ms Vinie Kouamou and all the supporting and administrative staff at Charles River Medical Group.

---

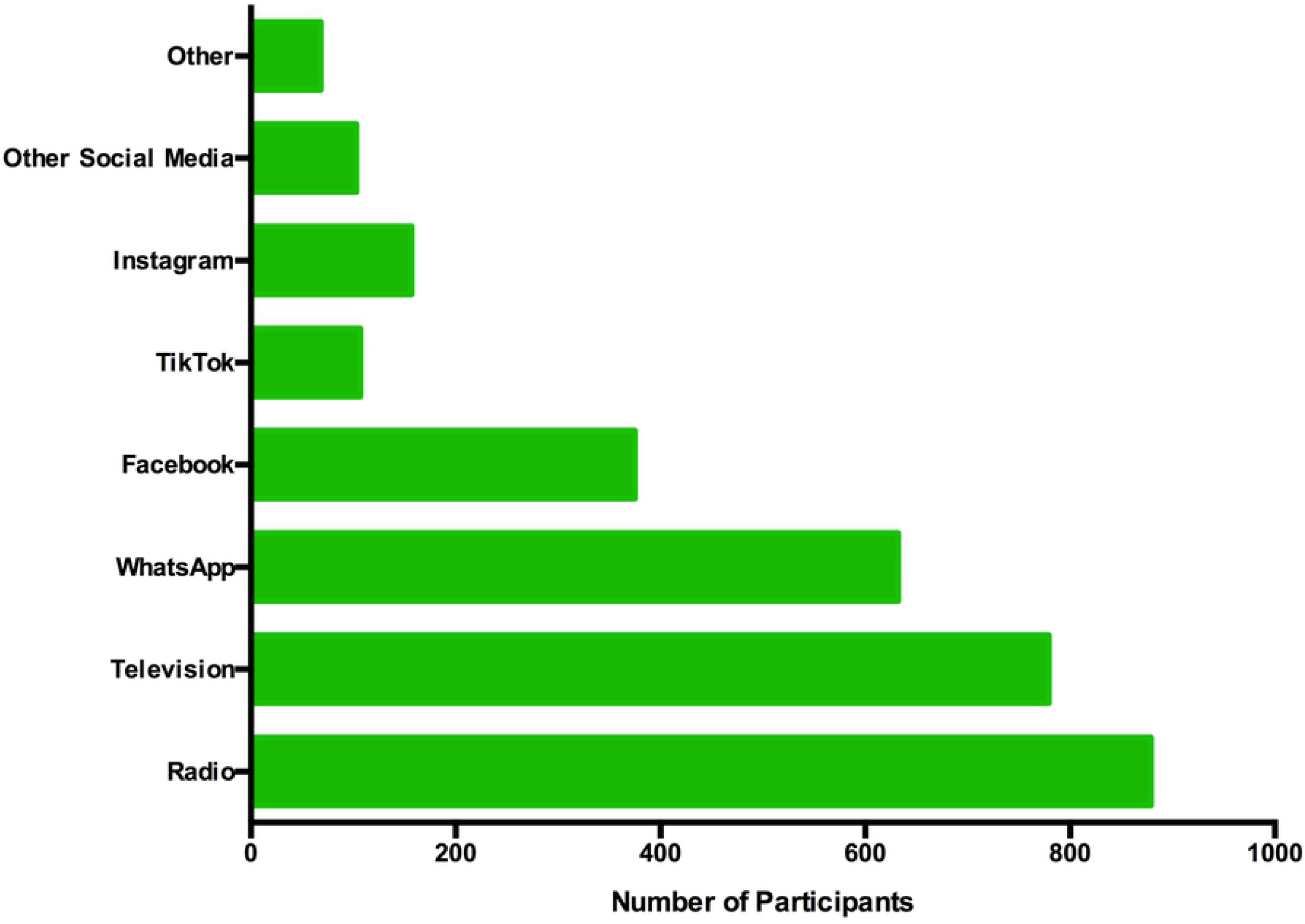

## Notes

### Competing Interest Statement

The authors have declared no competing interest.

### Funding Statement

Schmidt Futures Bill and Melinda Gates Foundation Funding to WJ, LS, SM, ATM The funders had no role in study design, data collection and analysis, decision to publish or preparation of the manuscript.

### Author Declarations

The study was reviewed and approved by the Medical Research Council of Zimbabwe ethics committee (MRCZ/A/2809). Written informed consent was obtained from all study participants.

